# Disparities in COVID-19 Hospitalizations and Mortality among Black and Hispanic Patients: Cross-Sectional Analysis from the Greater Houston Metropolitan Area

**DOI:** 10.1101/2020.08.19.20177956

**Authors:** Alan P. Pan, Osman Khan, Jennifer R. Meeks, Marc L. Boom, Faisal N. Masud, Julia D. Andrieni, Robert A. Phillips, Yordanos M. Tiruneh, Bita A. Kash, Farhaan S. Vahidy

**Author notes:** **Corresponding Author** Farhaan S. Vahidy, PhD MBBS MPH Josie Roberts Building, Suite 4.123 Houston Methodist 7550 Greenbriar Drive Houston, TX 77030.

## Abstract

**Background:** Disparate racial and ethnic burdens of the Coronavirus Disease 2019 (COVID-19) pandemic may be attributable to higher susceptibility to Severe Acute Respiratory Syndrome Coronavirus 2 (SARS-CoV-2) or to factors such as differences in hospitalization and care provision.

**Methods:** In our cross-sectional analysis of lab-confirmed COVID-19 cases from a tertiary, eight-hospital healthcare system (Houston Methodist) across greater Houston, multivariable logistic regression models were fitted to evaluate the odds of hospitalization and mortality for non-Hispanic Blacks (NHBs) vs. non-Hispanic Whites (NHWs) and Hispanics vs. non-Hispanics.

**Findings:** Between March 3^rd^ and July 18^th^, 2020, 70,496 individuals were tested for SARS-CoV-2; 12,084 (17·1%) tested positive, of whom 3,536 (29·3%) were hospitalized. Among positive cases, NHBs and Hispanics were significantly younger than NHWs and Hispanics, respectively (mean age NHBs vs. NHWs: 46.0 vs. 51.7 year and Hispanic vs. non-Hispanic: 44.0 vs. 48.7 years). Despite younger age, NHBs (vs. NHWs) had a higher prevalence of diabetes (25.2%), hypertension (47.7%), and chronic kidney disease (5.0%). Both minority groups resided in lower median income and higher population density areas. In fully adjusted models, NHBs and Hispanics had higher likelihoods of hospitalization, aOR (CI): 1·42 (1·24–1·63) and 1·61 (1·46–1·78), respectively. No differences were observed in intensive care unit (ICU) utilization or treatment parameters. Models adjusted for demographics, vital signs, laboratory parameters, hospital complications, and ICU admission demonstrated non-significantly lower likelihoods of in-hospital mortality among NHBs and Hispanics, aOR (CI): 0·65 (0·40–1·03) and 0·89 (0·59–1·31), respectively.

**Interpretation:** Our data did not demonstrate racial and ethnic differences in care provision and hospital outcomes. Higher susceptibility of racial and ethnic minorities to SARS-CoV-2 and subsequent hospitalization may be driven primarily by social determinants.

## Introduction

The Severe Acute Respiratory Syndrome Coronavirus 2 (SARS-CoV-2) pandemic is a global healthcare and economic catastrophe. The burden of the pandemic has been disparate. Elderly individuals and those with higher burden of cardio-metabolic diseases are in the highest risk category.^1,2^ Furthermore, reports have characterized varying levels of evidence suggesting that minority populations have been disproportionately impacted.^3,4^ However, there are relatively fewer reports that have evaluated an array of comprehensive socio-demographic, comorbidity, and clinical factors in establishing the association between race/ethnicity and likelihood of hospitalization and death due to COVID-19. Such reports are particularly lacking for Hispanic/Latino populations from healthcare systems that serve large, diverse populations. These knowledge gaps underscore the critical need to characterize the impact of the COVID-19 pandemic on minority populations in Texas.

At the time of this reporting, the greater Houston area has had over 118,000 confirmed cases of COVID-19 and upwards of 2,000 reported deaths.^5,6^ Houston has not only become a recent hotspot of the COVID-19 pandemic, but given the diverse makeup of its population, there is value in investigating how factors related to infection, critical illness, and outcomes are associated with race and ethnicity.

Houston Methodist (HM) is one of the largest healthcare systems in the greater Houston area. Our initial report characterized the racial and ethnic disparities seen with respect to SARS-CoV-2 susceptibility, noting higher susceptibility for SARS-CoV-2 positivity among non-Hispanic Black (NHB) and Hispanic populations, potentially mediated via population density and socio-economic status.^7^ In this study, we evaluate racial and ethnic differences in hospitalization and in-hospital mortality among the population testing positive for SARS-CoV-2. Given our prior data, we hypothesized that Black race and Hispanic ethnicity will be independently associated with a higher hospitalization rate and in-hospital mortality in the population of COVID-19 patients across HM.

## Methods

### Study Setting and Design

Houston Methodist (HM) comprises a flagship tertiary care hospital and seven community and continuing care hospitals across the greater Houston area. We retrospectively analyzed data from the COVID-19 Surveillance and Outcomes Registry (CURATOR) at HM. The CURATOR is an IRB-approved research registry designed to support COVID-19 research. The CURATOR data are populated from electronic health records and capture demographic, medical history, laboratory, medication, treatment, and outcomes variables for all individuals tested for SARS-CoV-2 at HM. Data are continually assessed for quality and are cross validated for accuracy.

Laboratory testing for SARS-CoV-2 was performed using nucleic acid amplification tests for qualitative detection of RNA from SARS-CoV-2 isolated and purified from nasopharyngeal specimens. Diagnostic panels for real-time reverse transcription polymerase chain reaction (RT-PCR) included the Panther Fusion® SARS-CoV-2 Assay and the Cepheid Xpert® Xpress SARS-CoV-2 Assay.

### Primary Exposure and Outcomes

Predefined categories of race and ethnicity were self-reported at the time of hospital encounter. Individuals identifying primarily as Black or African American were classified as “Black” and those identifying as Hispanic or Latino were categorized as “Hispanic”. The primary comparison categories for our study are non-Hispanic Blacks (NHBs) vs. non-Hispanic Whites (NHWs), and Hispanics vs. non-Hispanics. Individuals who had a hospital admission encounter with a laboratory confirmed COVID-19 positive test were classified as being hospitalized and among those who died during the episode of care related to COVID-19 were flagged for in-hospital mortality.

### Other Covariates

Other covariates include demographic characteristics such as age, sex, and insurance type. Population-level median income (inflation-adjusted to 2018 USD) was abstracted at the zip code tabulation area (ZCTA) level from the U.S. Census Bureau’s American Community Survey 5-year estimates (2014-2018).^8^ Land area estimates were obtained from the U.S. Census Bureau’s Gazetteer Files (2010).^9^ Population density (per square mile) was derived from population and land area estimates. Indices for socioeconomic status were set by designating zip codes (linked to ZCTA) representing the two lowest pentile groups for median income and the two highest pentile groups for population density.

Patient medical histories were queried for the presence of pre-existing conditions comprising the Charlson Comorbidity Index (CCI).^10^ Other conditions of interest include obesity, body mass index (BMI), hypertension, hyperlipidemia, asthma, smoking history, and prior vaccination history.

We extracted measurements of all systolic/diastolic blood pressure, respiratory rate, temperature, and oxygen saturation within the first six hours of admission (or the first six measurements). Laboratory parameters considered in our analysis include white blood cell count, blood differential (lymphocytes), platelet count, B-natriuretic peptide, procalcitonin, troponin, aspartate aminotransferase, alanine aminotransferase, C-reactive protein, ferritin level, D-dimer, creatinine, and venous lactate. Vital sign and laboratory measures were aggregated by taking the mean value for each patient, while excluding non-quantifiable results. Indicator flags characterizing abnormal results were derived based on commonly accepted clinical reference ranges. We excluded variables with data missingness ≥ 25% from multivariable models. Exact denominators are reported otherwise.

We extracted information on the development of hospital-associated complications (pneumonia, acute respiratory distress syndrome, bronchitis, lower respiratory tract infection, acute renal injury, acute hepatic injury, cardiomyopathy or congestive heart failure, hypoxic respiratory failure), utilization of high-acuity hospital resources such as admission to the Intensive Care Unit (ICU) and care involving use of invasive mechanical ventilation. The use of therapeutic agents such as Hydroxychloroquine, Ribavirin, Azithromycin, Lopinavir/Ritonavir, Remdesivir, Tocilizumab, Antithrombotics, Anticoagulants, and Dexamethasone were also evaluated.

### Statistical Analysis

Data are summarized as means (standard deviations), medians (interquartile range) or proportions based on the distribution and the functional form of the covariates. Bivariable analyses were performed to assess the associations between various sociodemographic, comorbidity and clinical factors by NHB vs. NHW race and Hispanic ethnicity. We present three explanatory logistic regression models each for the outcome of hospitalization and mortality, separately fitted for NHB (vs. NHW) race and Hispanic ethnicity (vs. non-Hispanic). This approach allowed us to adjust three distinct covariate sub-sets. For *hospitalization*: 1) Baseline models adjusted for age, sex, and insurance type, 2) Full socio-demographic models which included model-1 variables and added median household income and population density, and 3) Socio-demographic and comorbidity models which adds CCI, obesity, hypertension, and smoking to model-2. For *in-hospital mortality*: 1) Baseline socio-demographic and comorbidity models including age, sex, median household income, CCI, obesity, hypertension, and smoking, 2) Hospital admission models including model-1 and vital sign and laboratory measures, and 3) Comprehensive clinical models incorporating all variables of model-2 in addition to in-hospital complications, treatment course, and critical care utilization parameters.

We assessed the heterogeneity of our estimates across three strata for age (≤50, 51 – 75, and > 75 years) and time-period of hospitalization (Phase I: Up to April 15^th^, Phase II: April 16^th^ – May 31^st^, and Phase III: June 1^st^ – July 18^th^, 2020). All data preparation and analyses were performed using R statistical software (version 3·6·1; The R Foundation).

## Results

Between March 3^rd^ and July 18^th^, 2020, a total of 70,496 unique individuals were tested for SARS-CoV-2, of whom 12,084 (17·1%) tested positive. The mean age across the tested population was 49·9 years and 59·9% were females. Non-Hispanic Blacks (NHB) and Hispanics comprised 19·5% and 20·7% of the tested individuals respectively. Among laboratory confirmed COVID-19 cases, 3,536 (29·3%) were hospitalized, of whom 234 (6·6%) experienced in-hospital mortality. The proportions of tested, tested-positive, hospitalized and died individuals by minority race and ethnicity across Houston Methodist are presented in Figure 1.

**Figure 1.**
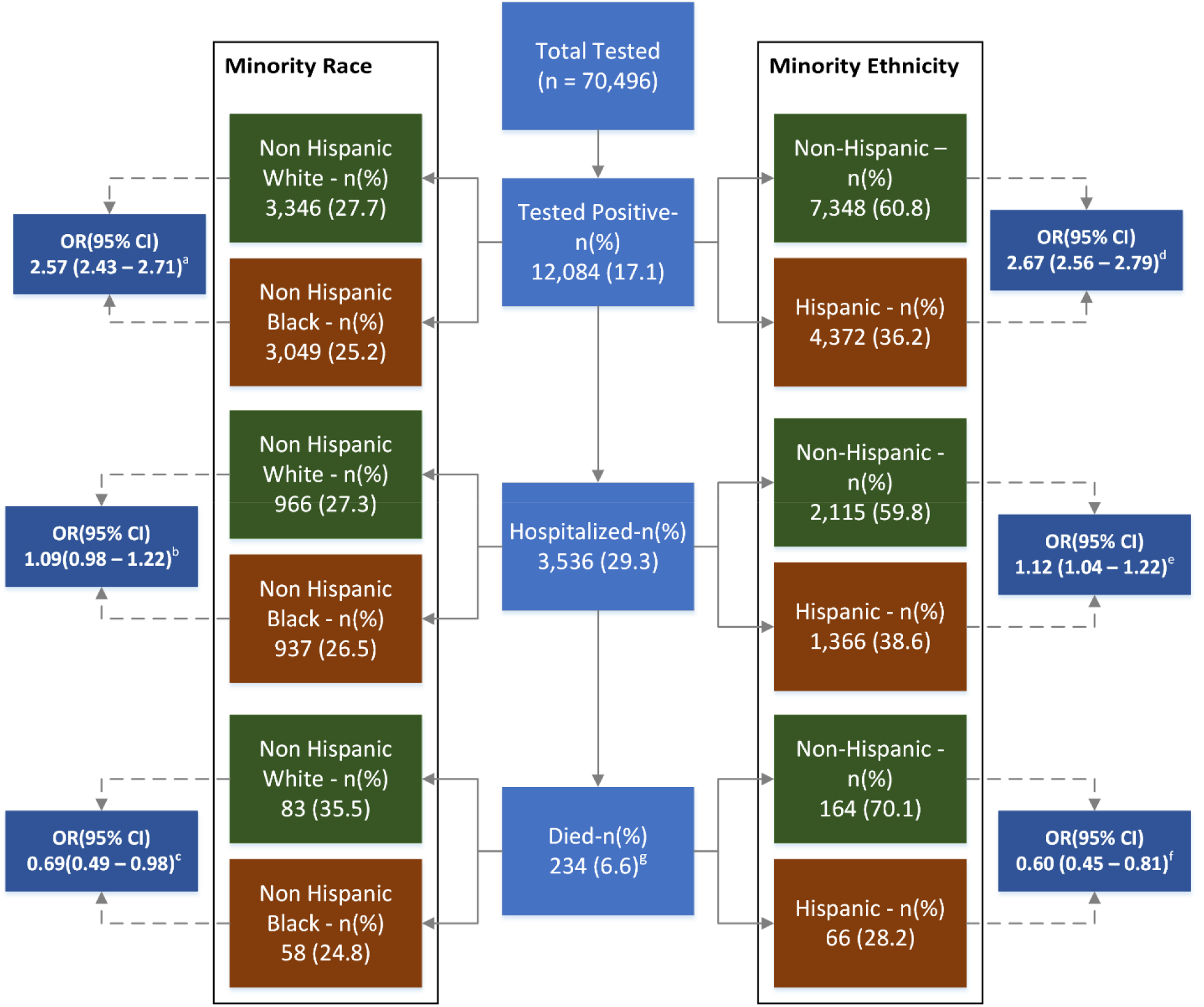
Frequency and proportions of SARS-CoV-2 and COVID-19 patients tested, hospitalized and died, by minority Race and Ethnicity. **Figure 1 Legend:** OR: Odds Ratio, 95% CI: 95% Confidence Interval, Respective ORs and 95% (CI) represent odds of testing positive, hospitalization and death for non-Hispanic Black (vs. non-Hispanic White) and Hispanic (vs. non-Hispanic).

### Baseline Socio-Demographic Characteristics of SARS-CoV-2 PCR-Confirmed Cases

Among the total confirmed cases, 3,049 (25·2%) identified as NHB and 4,372 (36·2%) were Hispanic. Compared to NHWs, NHBs were younger (46·0 vs. 51·7 years) with a higher proportion of females (60·2% vs. 53·6%). A greater proportion of NHBs resided in areas with lower income and higher population density. Despite younger age, NHBs (vs. NHWs) had a higher prevalence of diabetes (25·2% vs. 17·6%), hypertension (47·7% vs. 43·1%), kidney disease (5·0% vs. 3·3%) and AIDS (1·3% vs. 0·4%). Furthermore, a greater proportion of NHBs were current or former smokers (18·9% vs. 12·0%) and had a higher mean BMI (32·2 vs. 29·8 kg/m^2^). Other pre-existing conditions were more prevalent among NHWs, while the overall median Charlson Comorbidity Index (CCI) was observed to be lower for NHBs. A significantly smaller proportion of NHBs were previously vaccinated for influenza (56·9% vs. 67·7%) or pneumonia (37·8% vs. 49·5%).

Compared to non-Hispanics, Hispanics were younger (44·0 vs. 48·7 years) and had residence in lower income and higher population density areas. Hispanic SARS-CoV-2 positive individuals had lower prevalence of multiple pre-existing conditions, including hypertension, MI, CHF, PVD, COPD, and cancer. Overall comorbidity burden was also lower among Hispanics. Table 1 presents the baseline sociodemographic and comorbidity characteristics among SARS-CoV-2 PCR-confirmed cases, stratified by non-Hispanic race and Hispanic ethnicity.

**Table 1:** Socio-Demographic and Comorbidity Characteristics, by Race and Ethnicity, of Individuals who tested positive (n = 12,084) for SARS-CoV-2 at Houston Methodist through July 18^th^, 2020

|  | Non-Hispanic White<br>(n = 3,346) | Non-Hispanic Black<br>(n = 3,049) | Non-Hispanic<br>(n = 7,348) | Hispanic<br>(n = 4,372) |
| --- | --- | --- | --- | --- |
| **Demographics – n (%)** |  |  |  |  |
| Age – mean (SD) | 51.7 (20.1) | 46.0 (18.1) * | 48.7 (19.2) | 44.0 (17.4) * |
| Age ≤ 50 | 1582 (47.3) | 1825 (59.9) * | 4008 (54.5) | 3340 (76.4) * |
| Females | 1792 (53.6) | 1837 (60.2) * | 4164 (56.7) | 2398 (54.8) |
| Insurance Type |  |  |  |  |
| Commercial | 1736 (51.9) | 1454 (47.7) * | 3738 (50.9) | 1911 (43.7) * |
| Medicare | 892 (26.7) | 567 (18.6) | 1605 (21.8) | 574 (13.1) |
| Medicaid | 98 (2.9) | 227 (7.4) | 349 (4.7) | 380 (8.7) |
| Self-Pay | 507 (15.2) | 729 (23.9) | 1449 (19.7) | 1452 (33.2) |
| Other | 113 (3.4) | 72 (2.4) | 207 (2.8) | 55 (1.3) |
| Household Income, Median (IQR) | 75,793 (59,104 – 102,008) | 63,489 (46,801 – 76,163) * | 68,318 (52,825 – 97,681) | 59,104 (46,300 – 75,793) * |
| Population Density, Median (IQR) | 2741.6 (1162.5 – 4264.5) | 3256.8 (2040.7 – 4575) * | 2883.9 (1504 – 4384.8) | 3380.9 (1683.5 – 4384.8) * |
| **Coexisting Conditions / Prior Vaccinations – n (%)** |  |  |  |  |
| CCI Total, Median (IQR) | 2 (0 – 4) | 1 (0 – 3) * | 1 (0 – 4) | 1 (0 – 2) * |
| **Charlson Comorbidities** |  |  |  |  |
| Myocardial Infarction | 263 (7.9) | 224 (7.3) | 530 (7.2) | 156 (3.6) * |
| Congestive Heart Failure | 334 (10.0) | 328 (10.8) | 711 (9.7) | 202 (4.6) * |
| Peripheral Vascular Disease | 328 (9.8) | 200 (6.6) * | 570 (7.8) | 182 (4.2) * |
| CVA / TIA | 294 (8.8) | 259 (8.5) | 599 (8.2) | 186 (4.3) * |
| Dementia | 179 (5.3) | 115 (3.8) * | 317 (4.3) | 41 (0.9) * |
| COPD | 712 (21.3) | 589 (19.3) | 1409 (19.2) | 465 (10.6) * |
| Connective Tissue Disease | 91 (2.7) | 72 (2.4) | 177 (2.4) | 71 (1.6) * |
| Peptic Ulcer | 83 (2.5) | 51 (1.7) * | 147 (2.0) | 58 (1.3) * |
| Liver Disease (Mild) | 238 (7.1) | 123 (4.0) * | 410 (5.6) | 224 (5.1) |
| Liver Disease (Moderate to Severe) | 34 (1.0) | 12 (0.4) * | 48 (0.7) | 34 (0.8) |
| Diabetes w/o Complications | 590 (17.6) | 769 (25.2) * | 1541 (21.0) | 978 (22.4) |
| Diabetes with Complications | 199 (5.9) | 293 (9.6) * | 538 (7.3) | 219 (5.0) * |
| Hemiplegia | 40 (1.2) | 40 (1.3) | 87 (1.2) | 27 (0.6) * |
| CKD (Mild to Moderate) | 109 (3.3) | 152 (5.0) * | 283 (3.9) | 155 (3.5) |
| Solid Tumor (Localized) | 278 (8.3) | 131 (4.3) * | 438 (6.0) | 132 (3.0) * |
| Solid Tumor (Metastatic) | 214 (6.4) | 97 (3.2) * | 331 (4.5) | 99 (2.3) * |
| AIDS | 14 (0.4) | 41 (1.3) * | 56 (0.8) | 15 (0.3) * |
| BMI – mean (SD) | 29.8 (7.2) | 32.2 (8.2) * | 30.6 (7.7) | 31.7 (7.4) * |
| Obesity | 777 (23.2) | 721 (23.6) | 1614 (22.0) | 808 (18.5) * |
| Hypertension | 1441 (43.1) | 1454 (47.7) * | 3197 (43.5) | 1380 (31.6) * |
| Hyperlipidemia | 1093 (32.7) | 826 (27.1) * | 2182 (19.7) | 925 (21.2) * |
| Asthma | 374 (11.2) | 365 (12.0) | 811 (11.0) | 270 (6.2) * |
| Smoking (Current/Former) | 829 (12.0) | 577 (18.9) * | 1509 (20.5) | 607 (13.9) * |
| **Prior Vaccination<sup>†</sup>** |  |  |  |  |
| Influenza | 424/626 (67.7) | 370/650 (56.9) * | 861/1382 (62.3) | 322/544 (59.2) * |
| Pneumonia | 527/1064 (49.5) | 396/1047 (37.8) * | 996/2323 (42.9) | 353/1191 (29.6) * |

|  | Non-Hispanic<br>White<br>(n = 3,346) | Non-Hispanic<br>Black<br>(n = 3,049) | Non-Hispanic<br>(n = 7,348) | Hispanic<br>(n = 4,372) |
| --- | --- | --- | --- | --- |
| **Hospitalized** | 966 (28.9) | 937 (30.7) | 2115 (28.8) | 1366 (31.2) * |
| * $p < 0.05$<br>† Denominator denotes number of non-missing values<br>Excluding missing data:<br>Ethnicity (n=364)<br>Median Household Income (n=86)<br>Population Density (n=77)<br>CCI: Charlson Comorbidity Index, IQR: Interquartile Range |  |  |  |  |

### Racial and Ethnic Differences in Demographic, Clinical, and Outcome Characteristics Among Hospitalized COVID-19 Patients

Among all hospitalized COVID-19 patients (n = 3,536), NHWs and NHBs represented 27·3% and 26·5%, respectively, whereas non-Hispanics and Hispanics represented 59·8% and 38·6%, respectively. Sociodemographic and clinical differences among NHBs and NHWs and Hispanics and non-Hispanics are presented in Table 2.

**Table 2:** Clinical Characteristics of COVID-19 Hospitalized Patients (n = 3,536), by Race and Ethnicity, at Houston Methodist through July 18^th^, 2020

|  | Non-Hispanic<br>White<br>(n = 966) | Non-Hispanic<br>Black<br>(n = 937) | Non-Hispanic<br>(n = 2,115) | Hispanic<br>(n = 1,366) |
| --- | --- | --- | --- | --- |
| **Demographic Characteristics and Comorbidity Index – n (%)** |  |  |  |  |
| Age – mean (SD) | 65.0 (17.6) | 59.0 (16.4) * | 62.1 (17.3) | 54.2 (16.3) * |
| Female | 437 (45.2) | 513 (54.7) * | 1030 (48.7) | 642 (47.0) |
| Insurance Type |  |  |  |  |
| Commercial | 301 (31.2) | 332 (35.4) * | 718 (33.9) | 447 (32.7) * |
| Medicare | 478 (49.5) | 388 (41.4) | 956 (45.2) | 377 (27.6) |
| Medicaid | 27 (2.8) | 50 (5.3) | 82 (3.9) | 122 (8.9) |
| Self-Pay | 90 (9.3) | 122 (13.0) | 231 (10.9) | 386 (28.3) |
| Other | 70 (7.2) | 45 (4.8) | 128 (6.1) | 34 (2.5) |
| Household Income, Median (IQR) | 68,318 (56,974 – 94,829.3) | 56,974 (44,422 – 70,658) * | 65,805 (48,790 – 86,034) | 56,288 (44,422 – 70,324) * |
| Population Density, Median (IQR) | 2493.9 (812.2 – 4081.3) | 3256.8 (1925.8 – 4789.7) * | 2849.1 (1439.1 – 4304.3) | 3513.1 (1823.2 – 4988.2) * |
| CCI Total, Median (IQR) | 5 (2 – 8) | 3 (1 – 7) * | 4 (2 – 7) | 2 (1 – 4) * |
| **Vital Signs at Hospital Admission – n (%)** |  |  |  |  |
| SBP (mmHg) – mean (SD) | 133.8 (19.3) | 136.0 (20.8) * | 134.4 (19.9) | 131.7 (18.9) * |
| DBP (mmHg) – mean (SD) | 71.5 (9.3) | 74.1 (10.6) * | 72.6 (9.9) | 72.2 (9.3) |
| Respiratory Rate ≥ 24 breath / min | 172 (17.8) | 162 (17.3) | 384 (18.2) | 350 (25.6) * |
| Temperature ≥ 38°C | 31 (3.2) | 80 (8.5) * | 129 (6.1) | 89 (6.5) |
| Oxygen Saturation < 94% | 176 (18.2) | 111 (11.8) * | 335 (15.8) | 262 (19.2) * |
| **Laboratory Parameters – n (%)<sup>†</sup>** |  |  |  |  |
| WBC count <4000/μl | 79/964 (8.2) | 63/935 (6.7) | 159/2111 (7.5) | 74/1361 (5.4) * |
| Lymphocytes < 20% | 665/964 (69.0) | 554/934 (59.3) * | 1383/2110 (65.5) | 938/1360 (69.0) * |
| Platelet count <150,000/μl | 148/964 (15.4) | 110/935 (11.8) * | 284/2111 (13.5) | 145/1361 (10.7) * |
| B-natriuretic peptide >100 pg/ml, | 296/696 (42.5) | 213/646 (33.0) * | 566/1500 (37.7) | 245/954 (25.7) * |
| Procalcitonin >0.25 ng/ml | 149/319 (46.7) | 189/327 (57.8) * | 375/726 (51.7) | 248/526 (47.1) |
| Troponin ≥ 0.06 ng/ml | 206/607 (33.9) | 224/618 (36.2) | 475/1364 (34.8) | 186/633 (29.4) * |
| Aspartate aminotransferase > 40 U/l | 426/938 (45.4) | 467/919 (50.8) * | 1029/2068 (49.8) | 750/1337 (56.1) * |
| Alanine aminotransferase >40 U/l | 322/936 (34.4) | 317/917 (34.6) | 744/2064 (36.0) | 693/1335 (51.9) * |
| Total Bilirubin ≥ 1.2 mg/dl | 39/861 (4.5) | 43/865 (5.0) | 88/1930 (4.6) | 40/1254 (3.2) |
| C-reactive protein >8.2 ng/ml | 750/802 (93.5) | 725/777 (93.3) | 1661/1775 (93.6) | 1167/1199 (97.3) * |
| Ferritin level > 3000 ng/ml | 35/803 (4.4) | 70/774 (9.0) * | 114/1774 (6.4) | 65/1228 (5.3) |
| D-dimer > 0.5 ug/ml | 670/775 (86.5) | 681/766 (88.9) | 1511/1730 (87.3) | 985/1201 (82.0) * |
| Creatinine > 1.5 mg/dl | 154/956 (16.1) | 255/929 (27.4) * | 441/2097 (21.0) | 159/1346 (11.8) * |
| Venous Lactate > 2.2 mmol/l | 141/714 (19.7) | 122/669 (18.2) | 301/1557 (19.3) | 191/972 (19.7) |
| **Level of Hospital Care – n (%)** |  |  |  |  |
| ICU Stay – n (%) | 284 (29.4) | 297 (31.7) | 655 (31.0) | 432 (31.6) |
| On Ventilator – n (%) | 156 (16.1) | 162 (17.3) | 361 (17.1) | 212 (15.5) |
| Days on Ventilator, median (IQR)<sup>‡</sup> | 6.7 (2.5 – 13.3) | 6.9 (2.9 – 15.2) | 6.9 (2.8 – 14.8) | 8.0 (3.6 – 14.3) |
| **Hospital Complications – n (%)** |  |  |  |  |
| Pneumonia | 600 (62.1) | 617 (65.8) | 1366 (64.6) | 958 (70.1) * |
| ARDS | 35 (3.6) | 51 (5.4) | 109 (5.2) | 80 (5.9) |
| Bronchitis | 12 (1.2) | 14 (1.5) | 27 (1.3) | 16 (1.2) |
| Lower Respiratory Tract Infection | 12 (1.2) | 11 (1.2) | 24 (1.1) | 9 (0.7) |
| Acute Renal Injury | 229 (23.7) | 303 (32.3) * | 580 (27.4) | 191 (14.0) * |
| Acute Hepatic Injury | 16 (1.7) | 9 (1.0) | 28 (1.3) | 18 (1.3) |
| Cardiomyopathy or CHF | 151 (15.6) | 149 (15.9) | 321 (15.2) | 96 (7.0) * |
| Hypoxic Respiratory Failure | 347 (35.9) | 330 (35.2) | 764 (36.1) | 589 (43.1) * |
| **Therapeutics – n (%)** |  |  |  |  |
| Hydroxychloroquine | 109 (11.3) | 123 (13.1) | 258 (12.2) | 101 (7.4) * |
| Ribavirin | 33 (3.4) | 29 (3.1) | 71 (3.4) | 31 (2.3) |
| Azithromycin | 124 (12.8) | 150 (16.0) | 312 (14.8) | 208 (15.2) |
| Lopinavir/Ritonavir | 12 (1.2) | 8 (0.9) | 22 (1.0) | 2 (0.1) * |
| Remdesivir | 147 (15.2) | 139 (14.8) | 333 (15.7) | 345 (25.3) * |
| Tocilizumab | 146 (15.1) | 149 (15.9) | 354 (16.7) | 320 (23.4) * |
| Antithrombotic | 387 (40.1) | 367 (39.2) | 830 (39.2) | 329 (24.1) * |
| Anticoagulants | 874 (90.5) | 827 (88.3) | 1897 (89.7) | 1246 (91.2) |
| Dexamethasone | 389 (40.3) | 360 (38.4) | 841 (39.8) | 681 (49.9) * |
| **Outcomes – n (%)** |  |  |  |  |
| Currently Hospitalized | 182 (18.8) | 171 (18.2) | 391 (18.5) | 256 (18.7) |
| Died | 84 (8.7) | 58 (6.2) * | 164 (7.8) | 66 (4.8) * |
| Died (among discharged or died) | 84 (10.7) | 58 (7.6) * | 164 (9.5) | 66 (5.9) * |
| Died (ICU admits excl. currently hospitalized)<sup>§</sup> | 60/213 (28.2) | 44/209 (21.1) * | 124/478 (25.9) | 62/304 (20.4) * |
| Length of Stay Days – median (IQR) | 7 (3 – 15) | 6 (3 – 12) | 6 (3 – 13) | 5 (3 – 10) * |

\* $p < 0.05$
<sup>†</sup> Denominator denotes number of non-missing values
<sup>‡</sup> Non-zero ventilator days
<sup>§</sup> Denominator denotes number of ICU admits
<sup>||</sup> LOS only for discharged or died
Excluding missing data:
Ethnicity (n=55)
Median Household Income (n=34)
Population Density (n=30)
CCI: Charlson Comorbidity Index, IQR: Interquartile Range, SBP: Systolic Blood Pressure, DBP: Diastolic Blood Pressure, WBC: White Blood Cell Count, ARDS: Acute Respiratory Distress Syndrome, CHF: Congestive Heart Failure

NHBs were younger (59·0 vs. 65·0 years) with a higher percentage of females (54·7% vs. 45·2%), while NHWs had a higher overall risk burden (median CCI: 5 vs. 3). Similarly, Hispanics were younger (54·2 vs. 62·1 years), while non-Hispanics had a higher overall risk burden (median CCI: 4 vs. 2). Both minority groups were observed to reside in lower income and higher population density areas.

A greater proportion of NHBs (vs. NHWs) had mean admission temperature ≥ 38°C (11·8% vs. 3·2%), whereas a greater proportion of Hispanics (vs. non-Hispanics) had mean oxygen saturation < 94% (19·2% vs. 15·8%) and respiratory rate ≥ 24 breaths per min (25·6% vs. 18·2%). A greater proportion of NHBs had significantly elevated levels of procalcitonin (57·8% vs. 46·7%), aspartate aminotransferase (50·8% vs. 45·4%), ferritin (9·0% vs. 4·4%) and creatinine (27·4% vs. 16·1%). These laboratory parameters, along with higher respiratory rates and lower oxygen saturation, were also found to be associated with mortality in our bivariable analysis for factors associated with higher inhospital mortality (Appendix Table 1). Among Hispanics (vs. non-Hispanics), the proportion of patients with elevation of both liver enzymes (aspartate aminotransferase and alanine aminotransferase) was higher.

Although a greater proportion of NHBs (vs. NHWs) and Hispanic (vs. non-Hispanic) patients were admitted to the ICU, these differences were not statistically significant (NHBs vs. NHWs: 31·7% vs. 29·4% and Hispanic vs. non-Hispanic: 31·6% vs. 31·0%). Similarly, the differences in invasive mechanical ventilation utilization or days on mechanical ventilation did not reach statistical significance (Table 2).

A non-significantly higher proportion of NHBs (vs. NHWs) patients had pneumonia (65·8% vs. 62·1%); however, the proportion of Hispanic (vs. non-Hispanic) patients with in-hospital pneumonia was significantly higher (70·1% vs. 64·6%). This finding corresponded to a higher proportion of Hispanic (vs. non-Hispanic) patients developing hypoxic respiratory failure (43·1% vs. 36·1%). Among other complications, a significantly higher proportion of NHBs (vs. NHWs) demonstrated an acute renal injury (32·3% vs. 23·7%).

We did not observe significant differences in therapeutic usage among NHBs and NHWs. A significantly smaller proportion of Hispanics (vs. non-Hispanics) received hydroxychloroquine (7·4% vs. 12·2%), whereas the proportion of Hispanic patients receiving Remdesivir and Dexamethasone was significantly higher compared to non-Hispanics (Table 2).

Through July 18^th^, 2020, a greater proportion of NHBs vs. NHWs (30·7% vs. 28·9%) and Hispanics vs. non-Hispanics (31·2% vs. 28·8%) were hospitalized. Among discharged or deceased patients, the in-hospital mortality was significantly lower for both minority race/ethnicity groups compared with their respective counterparts (NHBs vs. NHWs: 7·6% vs. 10·7% and Hispanic vs. non-Hispanic: 5·9% vs. 9·5%). Similarly, lower proportional mortality for minority race/ethnicity was also observed among COVID-19 patients admitted to the ICU (NHBs vs. NHWs: 21·1% vs. 28·2% and Hispanic vs. non-Hispanic: 20·4% vs. 25·9%). The median hospital length of stay was shorter by one day for racial and ethnic minorities; this difference was statistically significant for Hispanic (vs. non-Hispanic) patients (Table 2).

### Racial and Ethnic Differences in Likelihood of Hospitalization Among SARS-CoV-2 Positive Patients

Our unadjusted analyses demonstrated a significantly higher likelihood of SARS-CoV-2 positivity among NHB (vs. NHW) (OR, 95% CI: 2·57, 2·45 – 2·71) and Hispanic (vs. non-Hispanic) (OR, 95% CI: 2·67, 2·56 – 2·79) race/ethnicity (Figure 1). Additionally, a higher likelihood of hospitalization was observed for NHB (vs. NHW) (OR, 95% CI: 1·09, 0·98 – 1·22) and Hispanic (vs. non-Hispanic) (OR, 95% CI: 1·12, 1·04 – 1·22) SARSCoV-2 positive patients (Figure 1). In all three multivariable models, NHB (vs. NHW) and Hispanic (vs. non-Hispanic) SARS-CoV-2 positive patients had a significantly higher likelihood of hospitalization (Table 3). In fully adjusted models, the aOR (95% CI) for NHBs (vs. NHWs) was 1·42 (1·24 – 1·63), whereas the aOR (95% CI) for Hispanics (vs. non-Hispanic) was 1·61 (1·46 – 1·78). Other factors independently associated with higher likelihood of hospitalization are additionally reported in Table 3. These factors also demonstrated an association with higher likelihood of hospitalization in the overall SARS-CoV-2 positive population (Appendix Table 2).

**Table 3:** Odds Ratios for Hospitalization among SARS-CoV-2 Positive Non-Hispanic Black and Non-Hispanic White Patients; and Hispanic and Non-Hispanic Patients

|  | Model 1<br>OR (95% CI)*<br>(n = 6,395) | Model 2<br>OR (95% CI)*<br>(n = 6,345) | Model 3<br>OR (95% CI)*<br>(n = 6,344) |
| --- | --- | --- | --- |
| Non-Hispanic Black (vs. NHW) | 1.74 (1.53, 1.98) | 1.61 (1.41, 1.84) | 1.42 (1.24, 1.63) |
| Age, in 5-year units | 1.30 (1.27, 1.34) | 1.31 (1.28, 1.34) | 1.18 (1.15, 1.22) |
| Female (vs. Male) | 0.64 (0.57, 0.72) | 0.63 (0.56, 0.71) | 0.63 (0.55, 0.71) |
| Insurance Type |  |  |  |
| Commercial | Reference | Reference | Reference |
| Medicare | 1.66 (1.39, 1.98) | 1.58 (1.32, 1.89) | 1.14 (0.94, 1.39) |
| Medicaid | 2.34 (1.72, 3.15) | 2.22 (1.63, 3.00) | 2.05 (1.48, 2.83) |
| Self-Pay | 0.98 (0.81, 1.17) | 0.94 (0.78, 1.13) | 1.25 (1.03, 1.52) |
| Residence: Low Income Zip Code<sup>†</sup> |  | 1.51 (1.32, 1.72) | 1.41 (1.23, 1.62) |
| Residence: Population Dense Zip<sup>‡</sup> |  | 0.89 (0.78, 1.01) | 0.96 (0.84, 1.10) |
| Charlson Comorbidity Index Score |  |  | 1.15 (1.12, 1.19) |
| Obesity |  |  | 1.50 (1.29, 1.74) |
| Hypertension |  |  | 1.77 (1.51, 2.07) |
| Smoking (Current/Former) |  |  | 0.94 (0.80, 1.10) |
|  | Model 1<br>OR (95% CI)*<br>(n = 11,720) | Model 2<br>OR (95% CI)*<br>(n = 11,645) | Model 3<br>OR (95% CI)*<br>(n = 11,642) |
| Hispanic (vs. Non-Hispanic) | 1.57 (1.43, 1.73) | 1.47 (1.33, 1.61) | 1.61 (1.46, 1.78) |
| Age, in 5-year units | 1.30 (1.28, 1.33) | 1.31 (1.29, 1.33) | 1.21 (1.18, 1.23) |
| Female (vs. Male) | 0.63 (0.57, 0.68) | 0.62 (0.56, 0.67) | 0.58 (0.53, 0.64) |
| Insurance Type |  |  |  |
| Commercial | Reference | Reference | Reference |
| Medicare | 1.70 (1.48, 1.95) | 1.63 (1.42, 1.87) | 1.23 (1.06, 1.42) |
| Medicaid | 3.05 (2.49, 3.72) | 2.88 (2.35, 3.52) | 2.80 (2.26, 3.46) |
| Self-Pay | 1.14 (1.02, 1.29) | 1.09 (0.97, 1.23) | 1.45 (1.28, 1.65) |
| Residence: Low Income Zip<sup>†</sup> |  | 1.56 (1.42, 1.72) | 1.48 (1.34, 1.64) |
| Residence: Population Dense Zip<sup>‡</sup> |  | 1.00 (0.91, 1.10) | 1.07 (0.97, 1.18) |
| Charlson Comorbidity Index Score |  |  | 1.12 (1.10, 1.15) |
| Obesity |  |  | 1.62 (1.45, 1.82) |
| Hypertension |  |  | 1.67 (1.49, 1.87) |
| Smoking (Current/Former) |  |  | 0.86 (0.77, 0.98) |
| * Adjusted Odds Ratios and 95% Confidence Intervals for association between individual co-variates and hospitalization among confirmed COVID-19 cases |  |  |  |
| † Low income classified as zip codes in the 2 lowest percentiles for median house income |  |  |  |
| ‡ High population density classified as zip codes in the 2 highest percentiles for population density |  |  |  |

### Racial and Ethnic Differences in Likelihood of Death Among Hospitalized COVID-19 Patients

The unadjusted likelihood of in-hospital mortality was significantly lower for NHB (vs. NHW) (OR, 95% CI: 0·69, 0·49 – 0·98) and for Hispanic (vs. non-Hispanic) (OR, 95% CI: 0·60, 0·45 – 0·81) COVID-19 patients. (Figure 1) Bivariable analysis of demographic and clinical factors associated with in-hospital mortality is presented in Appendix Table 1.

All multivariable models demonstrated a non-significantly lower likelihood of in-hospital mortality among NHB (vs. NHW) and Hispanic (vs. non-Hispanic) COVID-19 patients (Table 4). In fully adjusted models, the aOR (95% CI) of in-hospital mortality for NHBs (vs. NHWs) was 0·65 (0·40 – 1·03) and for Hispanics (vs. non-Hispanics) was 0·89 (0·59 – 1·31). Advanced age, hypoxemia, lymphopenia, thrombocytopenia, elevated hepatic enzymes and creatinine, development of pulmonary and extra-pulmonary complications, and ICU admission were all independently associated with in-hospital mortality. Patients receiving Remdesivir or Dexamethasone tended to have lower likelihood of mortality.

**Table 4:**
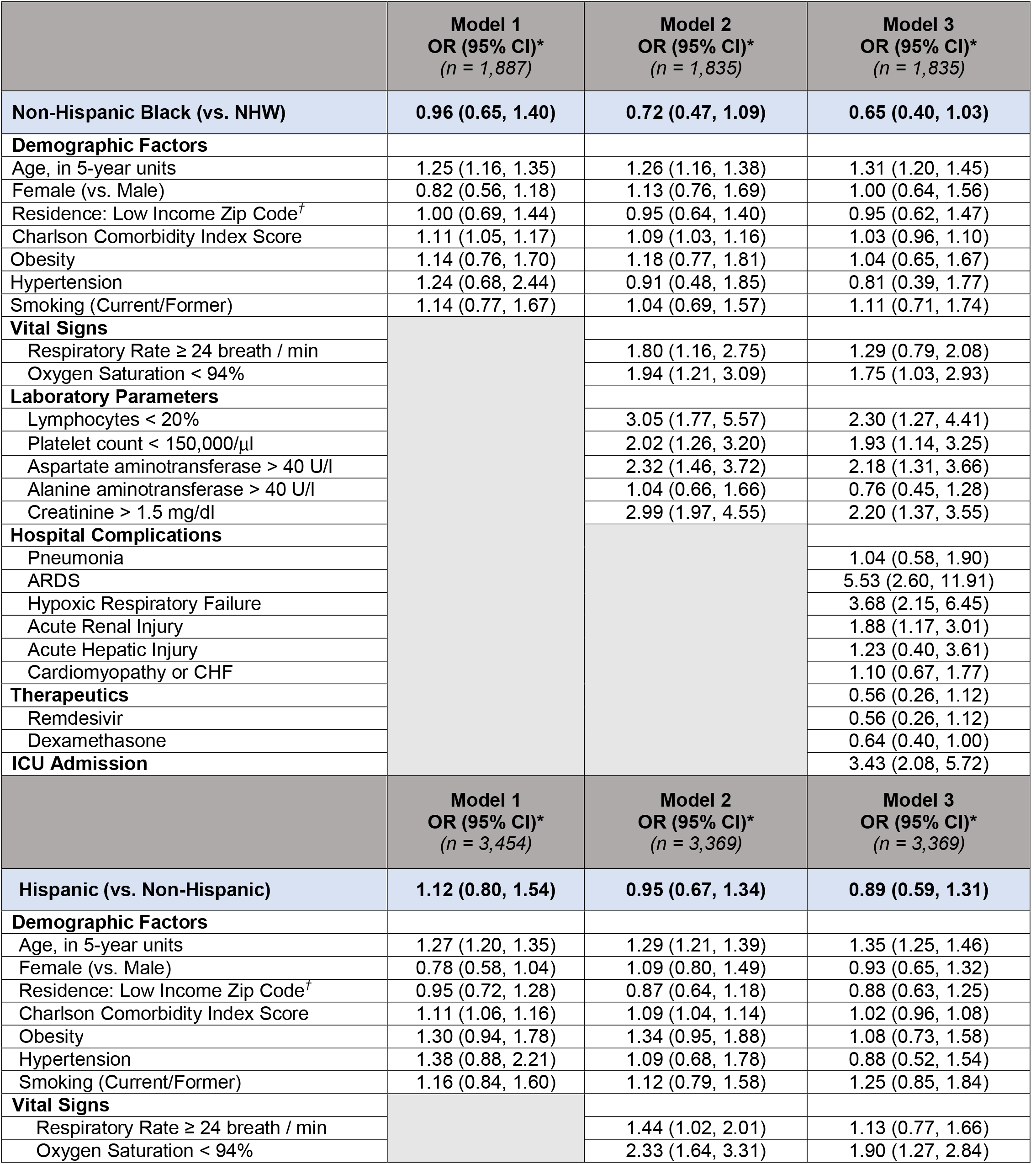
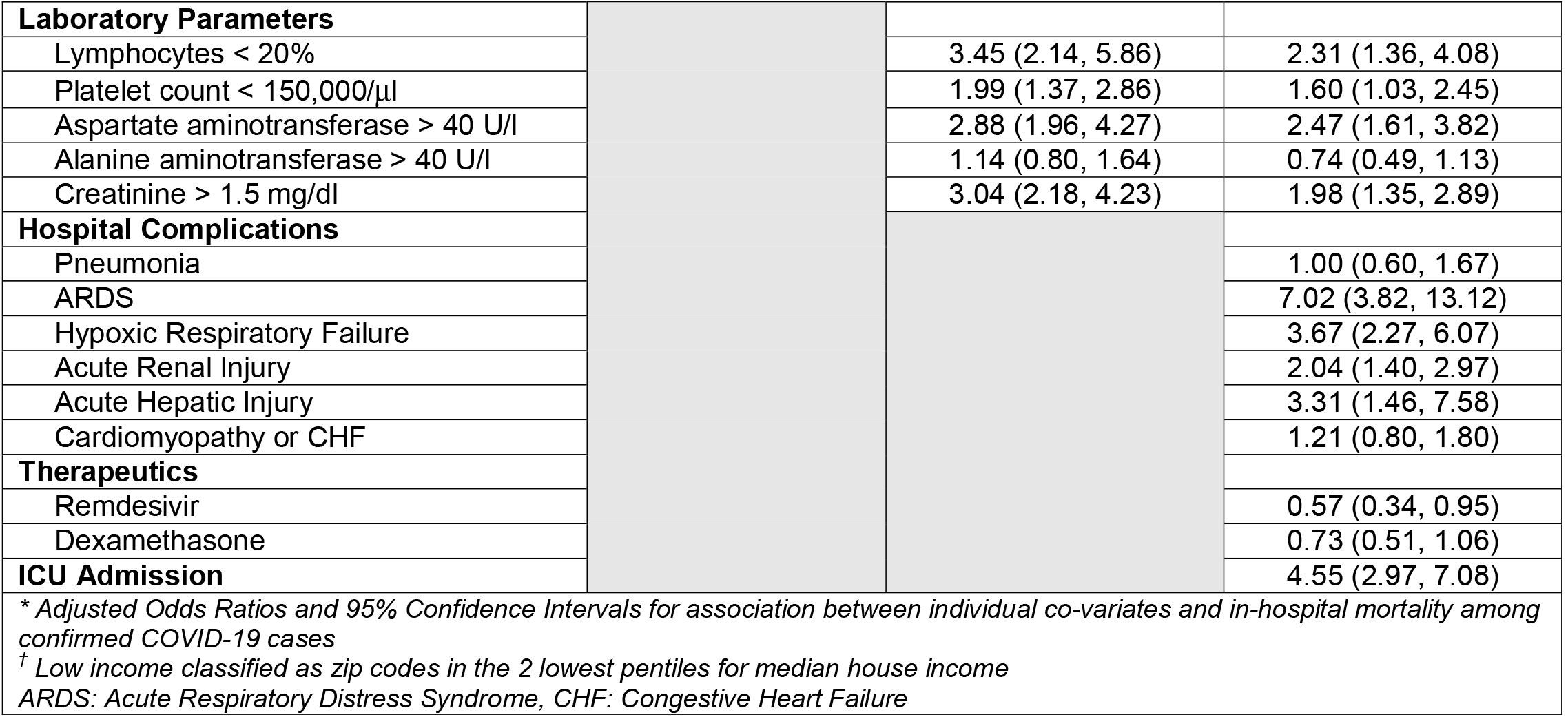
Odds for In-Hospital Mortality among Non-Hispanic Black and Non-Hispanic White Hospitalized COVID-19 patients

The evaluation of heterogeneity across the three strata of age and hospitalization phase did not yield significant interactions.

## Discussion

Initial reporting from the Houston Methodist (HM) system noted increased odds of SARS-CoV-2 positivity among non-Hispanic Blacks (NHBs) and Hispanics.^7^ Our current study expands on these findings and provides a comprehensive view of racial and ethnic disparities across varying stages in the progression of SARS-CoV-2 infectivity and COVID-19 outcomes. Among PCR-confirmed SARS-CoV-2 cases, NHBs and Hispanics were more likely to be hospitalized. No differences in hospital treatment were observed; however, and adjusted in-hospital mortality was non-significantly lower for minority race/ethnicity.

Detailed reports on racial and ethnic disparities from other large healthcare systems are generally limited; however, our findings are similar to those reported following an early investigation from the largest healthcare system in Louisiana which observed a higher likelihood of hospitalization among NHBs but no differences with respect to in-hospital mortality.^11^ Similarly, analyses from another single medical center in New York City reported no differences in hospital admission or in-hospital mortality for NHBs; however, Hispanics experienced increased likelihoods of both hospitalization and in-hospital mortality.^12^ Overall, higher susceptibility to infection and hospitalization has been broadly observed for minority groups – particularly Blacks^13,14,15,16^ and Hispanics^17^ – by a range of other studies, with fewer differences found with respect to mortality. It is therefore likely that the population-level burden of SARS-CoV-2 infection and COVID-19–related morbidity and mortality is driven primarily by a greater susceptibility to infection, rather than disparities related to care provision. It is critical to identify specific population-level targets for mitigating the disproportionate impact of the SARS-CoV-2 burden and similar health catastrophes among communities of color.

Interpretation of racial and ethnic disparities should also consider the distinct populations for which epidemiological analyses are being conducted. Whereas our hospital system data demonstrate a greater than twofold likelihood of SARS-CoV-2 infection among both racial and ethnic minority groups, publicly available data indicate that, in Houston, the population ratio of Black-to-White race is 0·91 but among the SARS-CoV-2 positive cases this ratio is 0·84 (demonstrating proportionately lower infection rates among Blacks) (Appendix Table 3). Conversely, the population ratio of Hispanic-to-non-Hispanic ethnicity is 0·81 but this ratio in the SARS-CoV-2 positive population is 0·85. Given this contrast between our findings and publicly reported data, it is important to note that the ratios for infection rates presented above reflect a snapshot of case prevalence. Our analyses do attempt to account for this when comparing the likelihoods of infection, hospitalization, or in-hospital mortality among our base cohorts. Furthermore, the differences may be explained, in part, by testing characteristics across a healthcare system; our data reflect characteristics of individuals who sought out point-of-care testing but do not capture the full scope of testing that is occurring at public or community-based sites across greater Houston. Presumably, we might expect differences in the location of preference for testing, based on an individual’s perception of need or convenience. In contrast to what we observed in Houston, the population Black-to-White ratio in New Orleans is 1·95, but the SARSCoV-2 infection and death Black-to-White ratios are 3·20 and 3·25, respectively. Similarly, higher Black-to-White ratios for SARS-CoV-2 infection, hospitalization and mortality (compared with the population Black-to-White ratio) were observed for New York City and Chicago. Appendix Table 3 presents available case and region demographics for a selection of major U.S. metropolitan areas. However, several gaps remain in reporting of race and ethnicity-based metrics.

Limitations of our study include use of data from a single healthcare system across the greater Houston area. Although HM serves patient populations in communities located beyond the metropolitan area, hospital data may not be representative of a generalized population. The findings from our data need to be replicated across large and diverse healthcare systems and evaluated in light of specific demographic contexts in the region. Additionally, given the observational nature of our study, all estimates are associative and do not imply causation. In particular, we did not perform adjusted analyses for factors associated with hospitalization or mortality, as that was not the focus of this study. Any association observed in our bivariable analysis of factors related to morality or hospitalization would warrant separate validation analyses to be of interpretational value. Median household income and population density were included as approximations of socioeconomic and social distancing challenges; however, the true effects are undoubtedly more complex and multifactorial in nature.^18,19^ Future analyses should employ additional population measures to account for social determinants of health.

## Conclusion

The need to identify the root causes of the disparate burden of the SARS-CoV-2 pandemic among communities of color as precisely as possible should lead healthcare officials and institutions to focus the public health messaging on appropriate targets to mitigate such disparities. Our data demonstrate that, despite a higher likelihood of infection and hospitalization among NHB and Hispanic populations, there were no subsequent differences in hospital treatment or in-hospital mortality. These findings support the hypothesis that the overall higher burden of the COVID-19 pandemic among racial and ethnic minorities may be driven by higher susceptibility to contracting the SARS-CoV-2. Such susceptibilities are most likely mediated by adverse social determinants of health, which puts these minority populations at risk of infection in the community setting.

## Data Availability

All data sharing and collaboration requests should be directed to the corresponding author.

## Data sharing

All data sharing and collaboration requests should be directed to the corresponding author.

